## Supplementary material for "Development of a novel startle response task in Duchenne muscular dystrophy": S1 Table

| | Gao *et al*. (2010)[1] | Pattwell *et al*. (2012)[2] | Neumann *et al*. (2008)[3] | Shechner *et al*. (2015)[4] | Field *et al*. (2006)[5] | Lau *et al*. (2008)[6] | Glenn *et al*. (2012)[7] | Jovanovic *et al*. (2014)[8] | Schiele *et al*. (2016)[9] |
|---|---|---|---|---|---|---|---|---|---|
| **Subjects** | | | | | | | | | |
| **Age range** | 3-8 years | 5 - 28 years | 8-17 years | youth/adults | | | young adults | 8-13 years | 8-10; 18-50 |
| **Total number** | 200 | | 16 | youth: 37 adults: 47 | | 54 | 40 | 60 | children: 239 adults: 278 |
| **Healthy/ pathology** | Healthy | Healthy | Healthy | Healthy/ anxious | Healthy | Healthy (38)/ anxious (16) | | Low/high anxiety | Healthy |
| **Conditioned stimulus (CS)** | | | | | | | | | |
| **CS+/CS-** | 1000Hz/500Hz 60dB tone | Coloured squares | Black & white squares | Coloured bell pictures | Neutral cartoons | Neutral faces | Neutral faces & shapes | Coloured shapes | Neutral faces |
| **Duration (s)** | 12.5 | 3 | 8 | 8 | 3 | 8 | 6 | 0.50 | 6 |
| **Unconditioned stimulus (UCS)** | | | | | | | | | |
| **UCS** | White noise + rattling keys | White noise + tone | Metal on slate sound | Alarm & red bell pic | Food pictures | Fearful face + scream | Fearful face + scream/shock | Noise burst | Fearful face and scream |
| **Duration (s)** | 4.5 | 1 | 3 | 1 | 2 | 3 | 3 (face); 1 (scream) | 0.04 | 1.5 |
| **Onset (s)** | 10 | 2 | 5 | 7 | 1 | at CS+ offset | at CS+ offset | same time | at CS+ offset |
| **Offset (s)** | 2s after CS+ | with CS+ | with CS+ | with CS+ | with CS+ | | | | |
| **dB level** | 90 | 85-95 (child); 94-104 (adol/adult) | 83 | 95 | n/a | 95 | 80 | 106 | 95 |
| **Mean inter-trial interval (ITI) (s)** | 38 | 13 | 14.5 | 14.5 | 3 | | 11 | 15.5 | 10.5 |
| **ITI range (s)** | 34-42 | 13 | 13-16 | 8-21 | 2-4 | | 10-12 | 9-22 | 9-12 |
| **Analysis window** | | 1-10s | | 0-5s | | | | 3-6s | |
| **Trial protocol** | | | | | | | | | |
| **Orienting/pre-exposure** | 6 neutral tones | | 3 min baseline | 6 startle probes | | | | | |
| **No. Familiarisation trials** | | | 2 CS+/2CS- | 4 CS+/4 CS- | | 4 CS+/4 CS- | | 0 | 4 CS+/4 CS- |
| **No. Acquisition trials** | | | | | | | | | |
| **CS+ (reinforced)** | 6 | 12 | 12 | 8 | 20 | 12 | 6 | 3 | 10 |
| **CS+ (not reinforced)** | 3 | 12 | 0 | 2 | 0 | 4 | 2 | 3 | 2 |
| **CS-** | 3 | 24 | 12 | 10 | 4 | 16 | 8 | 3 | 12 |
| **No. Extinction trials:** | n/a | 24 CS+/24 CS- | 12 CS+/12 CS- | 8 CS+/8 CS- | 20 CS+ | 15 CS+/15 CS- | | 12 CS+ | n/a |
| **Counter-balanced CS** | N | Y | Y | Y | N | N | Y | N | |
