## Supplementary material for "Development of a novel startle response task in Duchenne muscular dystrophy": STROBE Statement

*N.B. There is not currently a specific checklist for methodological studies in health research, therefore the STROBE checklist has been used although not all sections are completely applicable. A new checklist, the MISTIC (MethodologIcal STudy reportIng Checklist) is currently under development: https://bmjopen.bmj.com/content/10/12/e040478.full*

| | Item No | Recommendation | Page No |
|---|---|---|---|
| **Title and abstract** | 1 | (*a*) Indicate the study's design with a commonly used term in the title or the abstract | 2 |
| | | (*b*) Provide in the abstract an informative and balanced summary of what was done and what was found | 2 |
| **Introduction** | | | |
| Background/rationale | 2 | Explain the scientific background and rationale for the investigation being reported | 4-9 |
| Objectives | 3 | State specific objectives, including any prespecified hypotheses | 7-9 |
| **Methods** | | | |
| Study design | 4 | Present key elements of study design early in the paper | 9-12 |
| Setting | 5 | Describe the setting, locations, and relevant dates, including periods of recruitment, exposure, follow-up, and data collection | 9 |
| Participants | 6 | (*a*) Give the eligibility criteria, and the sources and methods of selection of participants | 9 |
| Variables | 7 | Clearly define all outcomes, exposures, predictors, potential confounders, and effect modifiers. Give diagnostic criteria, if applicable | 9-10, 14 |
| Data sources/ measurement | 8* | For each variable of interest, give sources of data and details of methods of assessment (measurement). Describe comparability of assessment methods if there is more than one group | 12-15 |
| Bias | 9 | Describe any efforts to address potential sources of bias | 10 |
| Study size | 10 | Explain how the study size was arrived at | 21 |
| Quantitative variables | 11 | Explain how quantitative variables were handled in the analyses. If applicable, describe which groupings were chosen and why | 21-22 |
| Statistical methods | 12 | (*a*) Describe all statistical methods, including those used to control for confounding | 22-23 |
| | | (*b*) Describe any methods used to examine subgroups and interactions | n/a |
| | | (*c*) Explain how missing data were addressed | 14-17 |
| | | (*d*) If applicable, describe analytical methods taking account of sampling strategy | n/a |

| | | | |
|---|---|---|---|
| | | (*e*) Describe any sensitivity analyses | n/a |
| | | **Results** | |
| Participants | 13* | (a) Report numbers of individuals at each stage of study—eg numbers potentially eligible, examined for eligibility, confirmed eligible, included in the study, completing follow-up, and analysed | 23-24 |
| | | (b) Give reasons for non-participation at each stage | 24 |
| | | (c) Consider use of a flow diagram | n/a |
| Descriptive data | 14* | (a) Give characteristics of study participants (eg demographic, clinical, social) and information on exposures and potential confounders | 24 |
| | | (b) Indicate number of participants with missing data for each variable of interest | 24 |
| Outcome data | 15* | Report numbers of outcome events or summary measures | 25-26 |
| Main results | 16 | (*a*) Give unadjusted estimates and, if applicable, confounder-adjusted estimates and their precision (eg, 95% confidence interval). Make clear which confounders were adjusted for and why they were included | 25-26 |
| | | (*b*) Report category boundaries when continuous variables were categorized | n/a |
| | | (*c*) If relevant, consider translating estimates of relative risk into absolute risk for a meaningful time period | n/a |
| Other analyses | 17 | Report other analyses done—eg analyses of subgroups and interactions, and sensitivity analyses | 26-30 |
| | | **Discussion** | |
| Key results | 18 | Summarise key results with reference to study objectives | 30-31 |
| Limitations | 19 | Discuss limitations of the study, taking into account sources of potential bias or imprecision. Discuss both direction and magnitude of any potential bias | 32 |
| Interpretation | 20 | Give a cautious overall interpretation of results considering objectives, limitations, multiplicity of analyses, results from similar studies, and other relevant evidence | 32 |
| Generalisability | 21 | Discuss the generalisability (external validity) of the study results | n/a |
| | | **Other information** | |
| Funding | 22 | Give the source of funding and the role of the funders for the present study and, if applicable, for the original study on which the present article is based | Financial Disclosure Statement |

*Give information separately for exposed and unexposed groups – *n/a*
